# DIASTOLOGY AND MITRACLIP OUTCOMES: MULTICENTER REAL WORLD EVIDENCE STUDY

**DOI:** 10.1101/2025.04.25.25326462

**Authors:** Vivek Joseph Varughese, Chandler Richardson, James Pollock, Patryk Czyzewski, Hata Mujadzic, Michael Cryer

## Abstract

**INTRODUCTION:** MitraClip^®^ (MC) placement has been extensively used as an intervention for mitral transcatheter edge-to-edge repair (mTEER) for functional mitral regurgitation (FMR). The aim of our study is to analyze the association between pre procedural echocardiographic parameters of diastolic function (DF) and one-year outcomes after MC placement.

**STUDY DESIGN AND METHODS:** The study was designed in retrospective longitudinal cross-sectional format. 224 patients who underwent MC placement between January of 2021 and March of 2024 at Prisma Health Richland, Baptist and Greenville were included in the study. Primary Efficacy Endpoint (PEE) was determined by absence of heart failure hospitalizations requiring Intravenous Diuretics or Cardiac related death in the one year follow up. One-way ANOVA was used to determine variance of echo parameters between groups. Multivariate regression analysis was done to identify pre-procedural echocardiographic parameters of DF that held significant association with failure to reach the PEE. A two tailed p-value < 0.05 was used to determine statistical significance.

**RESULTS:** Of the 224 patients included in the study, 85 patients (37.94%) failed to reach the PEE or had symptom worsening. The mean mitral valve (MV) deceleration time was 176.88 ms (164.14 - 189.62) in the symptom worsening group compared to 201.53 ms (186.01 - 217.07) in the symptom improvement group. Mean of E/A ratio (MV peak E velocity/ A velocity) was noted to be 2.35 (1.97 - 2.74) in the symptom worsening group compared to 1.90 (1.68 - 2.13) in the symptom improvement group. After multivariate regression analysis, E/A ratio was found to have significant association with failure to reach PEE: Odds Ratio (OR): 1.61 (1.13 - 2.29), p-value: 0.008. Area under the curve (AUC) analysis for the E/A ratio was calculated at 0.603 (0.50 - 0.69) for symptom worsening group.

**CONCLUSIONS:** Patients that failed to reach the PEE had a lower pre-procedural MV deceleration time of 176.88 ms (164.14 - 189.62), however no association was observed between MV deceleration time and failure to reach the PEE in the multivariate regression analysis. Pre-procedural E/A ratio had significant association with symptom worsening after multivariate regression analysis: OR: 1.61 (1.13 - 2.29). AUC for E/A ratio in symptom worsening group was 0.603, making it a moderate predictor than random guessing for failure to reach the PEE.

## INTRODUCTION

Chronic mitral regurgitation (MR) can be broadly subdivided into primary and secondary mitral regurgitation. Primary mitral regurgitation (PMR) occurs from defects associated with the mitral valve apparatus, i.e., the leaflets, annulus, papillary muscles or chordae tendineae. Functional mitral regurgitation (FMR) stems from defects in coaptation of the mitral leaflets due to cardiac remodeling leading to an abnormal papillary muscle interaction with the leaflets, not necessarily due to anatomic defects of the mitral valve (MV) apparatus. This can be from wall motion abnormalities and papillary muscle dysfunction related to prior myocardial infarction, cardiac remodeling related to long-standing heart failure, and/or concomitant left atrial dilatation most commonly from long-standing atrial fibrillation [1]. The difference they hold in etiology also reflects in the management strategies as well as the long-term outcomes. PMR is essentially a structural defect of the mitral valve apparatus, and management relies on the correction, often surgical, of the valve apparatus, while FMR behaves more like a consequence of cardiac remodeling.

Looking at the current AHA/ACC guidelines on the management of valvular heart diseases, FMR is recommended for structural heart team evaluation, with a heart failure specialist being an integral part of the team. FMR can be reversible through remodeling achieved through guideline directed medical therapy (GDMT), revascularization, and resynchronization strategies. Regurgitant Fraction > 50%, Regurgitant Volume > 60mL, Vena Contracta ≥ 0.7cm and Effective Regurgitant Orifice Area (EROA) ≥ 0.4 cm^2^ are the echocardiographic parameters defining severity in both PMR and FMR [2]. In symptomatic severe PMR, as well as asymptomatic severe PMR with evidence of LV dysfunction (defined as Ejection fraction < 60% or LVEDD ≥ 4 cm), recommendations are for mitral valve surgery or repair with mitral transcatheter edge-to-edge repair (mTEER) recommended only in high or prohibitive-risk surgical candidates [2]. GDMT holds a role in patients who are not candidates for interventions or while waiting for the intervention. While in the management of FMR, interventions like valvular replacement, repair or mTEER are recommended in severe MR with persistent symptoms despite optimal GDMT and revascularization strategies [2].

Mitral transcatheter edge-to-edge repair (mTEER) is a transcatheter approach to mechanically fix the malcoaptation of mitral leaflets caused due to annular dilatation in FMR. Current guidelines recommend it as a treatment modality in severe symptomatic FMR with persistent symptoms despite maximally tolerated GDMT, ejection fraction (EF) between 20% and 50%, given the pulmonary artery systolic pressure (PASP) remains less than 70 mm Hg and left ventricular end systolic dimension (LVESD) remains less than 7 cm [2]. Surgical mitral valve repair or replacement is recommended in patients who are not candidates for mTEER [2]. This puts MitraClip^®^ as more of a management strategy for advanced heart failure rather than a structural intervention for a primary valve disease.

FMR is a consequence of cardiac remodeling due to advanced heart failure, at the same time, is also a factor worsening cardiac remodeling, creating a vicious cycle. This makes the management challenging, often creating a gray area between the switch from medical management to interventions. This is analogous to the situation of ischemic cardiomyopathy management, where trials like the STITCH compared revascularization strategies (CABG) to medical management, and COURAGE trial, that compared to Percutaneous Coronary Intervention (PCI) with GDMT to GDMT alone in stable ischemic heart disease [3,4]. COAPT and the MITRA FR were two of the landmark clinical trials that studied outcomes for mTEER [5,6]. While the COAPT [5] trial demonstrated better outcomes in terms of heart failure hospitalization and death in 24 months in the mTEER plus GDMT group compared to GDMT alone, the MITRA FR trial showed no difference in outcomes between the groups. However, the extent of cardiac remodeling in patients who were included in the MITRA FR trial was substantially higher, including patients with LVESD > 7cm and a lower mean effective regurgitant orifice area (EROA) (0.3 cm^2^) compared to patients included in the COAPT trial (0.4 cm^2^) [5,6]. RESHAPE-HF2 was a later trial that proved superior outcomes in the mTEER plus GDMT group in terms of one year heart failure hospitalizations and death, as well as improvement from the baseline Kansas City Cardiomyopathy Questionnaire (KCCQ) scores [7].

With advanced heart failure strategies, including assist devices, continuous inotropic infusions and heart transplantations becoming more robust and prevalent, selection of patients to have a meaningful difference in heart failure symptoms through mTEER becomes relevant. Insights from the COAPT and MITRA FR trials gave rise to the disproportionate MR theory [8], and the predictive value of the EROA to the LVEDV (Left Ventricular End Diastolic Volume) ratio have been studied in various sub trials of the COAPT. However, the crescentic nature of the regurgitant orifice in FMR, leading to eccentric regurgitant jets would lead to underestimation of the EROA by proximal isovelocity surface area (PISA) method. 3D models and use of vena contracta area are being studied for the effective quantification of the regurgitation [9].

There have been retrospective studies in the past studying global longitudinal strain post procedure as an outcome predictor after mTEER, but there is a general paucity in data analyzing the impact of diastolic function in FMR [10]. Possible explanations include the overestimation of the MV peak E velocity due to higher trans-mitral pressure gradient created due to the regurgitant jet, a lower A velocity and therefore overestimation of the E/A ratio, which becomes important in analyzing diastolic function in reduced ejection fraction states [5]. However, irrespective of the regurgitation severity, pulmonary vascular congestion due to elevated end diastolic pressures is the primary symptom driver in FMR. In this context, the extent of diastolic dysfunction can have a potential impact on meaningful symptomatic improvement with mTEER as LA and LV compliance can have major effects on end diastolic pressures. The aim of our study is to analyze the association between pre -procedural echocardiographic parameters of LV diastolic function and one-year outcomes after MitraClip^®^ placement.

## STUDY DESIGN AND METHODS

### STUDY DESIGN

The study was designed in a retrospective longitudinal cross-sectional format. Patients who underwent mTEER using MitraClip^®^ were selected for the study. Patients who underwent the procedure at Prisma Health Richland Hospital, Prisma Health Baptist Hospital and Prisma Health Greenville campus between January of 2021 and March of 2024 were selected for the study.

Inclusion Criteria: Patient aged > 18, who underwent MitraClip^®^ placement for SMR, also known as functional mitral regurgitation (FMR), with a 3+/4+ severity of mitral regurgitation.

Exclusion Criteria: Patient undergoing MitraClip^®^ placement for PMR. Patients without one-year surveillance echocardiogram and KCCQ score documentations were excluded from the study. Patients with coexisting diagnosis of terminal illness like malignancies, and patient death in the one-year follow up period due to non-cardiovascular related conditions were excluded from the study. Patients with advanced heart failure treatment options like LVAD in the one-year follow up were excluded from the study. Patients requiring repeat clip placements/ required surgical valve repair in the one year follow up were excluded. Patients with comorbid mitral stenosis were excluded from the study.

After careful manual chart review of 326 patients, 224 patients were included in the study. One-year outcomes were analyzed for the selected patients according to the criteria defined by the Mitral Valve Research Consortium on efficacy and safety endpoints.

As per the recommendations of the mitral valve research consortium, **Primary efficacy endpoint** was defined by absence of hospitalizations for heart failure/ volume overload in the one year following mTEER placement requiring IV diuretics or cardiovascular related deaths in the one-year post MitraClip^®^ placement. **Safety endpoints** were also defined as per the criteria specified by the mitral valve research consortium defined as ER/ hospital visits in the 30 days following mTEER placement for diagnoses that could be attributed to the procedure.

### Study subjects were divided into three groups

1. ymptom Improvement Group (Group 1): patients who had a change in their baseline KCCQ (Kansas City Cardiomyopathy Questionnaire) score > 20 at one year and absence of heart failure hospitalization/ death and <50% increase in their preprocedural diuretic usage.
2. Stable Symptom Group (Group 2): patients with no heart failure hospitalization or death in the one year follow up period, but with a change in their baseline KCCQ score < 20 or increase in their diuretic usage > 50%.
3. Symptom worsening group (Group 3): patients who had heart failure related hospitalizations or death in the one year follow period

### METHODOLOGY AND STATISTICS

Study subjects were divided into three groups as per the one-year outcomes specified in the study design. First part of the statistical approach was determining the pre- and post-procedural echocardiographic dimensions indicative of the LV diastolic function, with corresponding changes. Normality of each variable was assessed by using the Kolmogorov-Smirnov test. Quantitative data was expressed by mean, standard deviation and difference between means of two groups were tested by Mann Whitney U test while for more than two groups comparison Kruskal Wallis H test was used. Univariate and multivariate logistic regression analysis was done to identify factors affecting symptom worsening and adjusted odds ratio was calculated along with 95% CI. ‘P’ value less than 0.05 was considered statistically significant. Age, sex, race, preprocedural KCCQ score, Left Ventricular End Systolic Volume (LVESV), Left Ventricular End Diastolic Volume (LVEDV), Effective Regurgitant Orifice Area (EROA), Pulmonary Artery Systolic Pressure (PASP), history of atrial fibrillation and adherence with medications/revascularization strategies in the follow up period (ACE/ARB, beta blockers/ cardiac resynchronization therapy) were used in the multivariate regression analysis. LVEDV, LVESV, EROA and PASP are echocardiographic parameters with proven association with outcomes after Mitra clip placement.

## RESULTS

At One year follow up, 94 patients had symptom improvement, 45 patients were symptom stable, and 85 patients had symptom worsening/death.

Pre-procedural Echocardiographic parameters between the groups were compared, and variance analyzed across the groups. Results are summarized in Table 1.

**TABLE 1.**
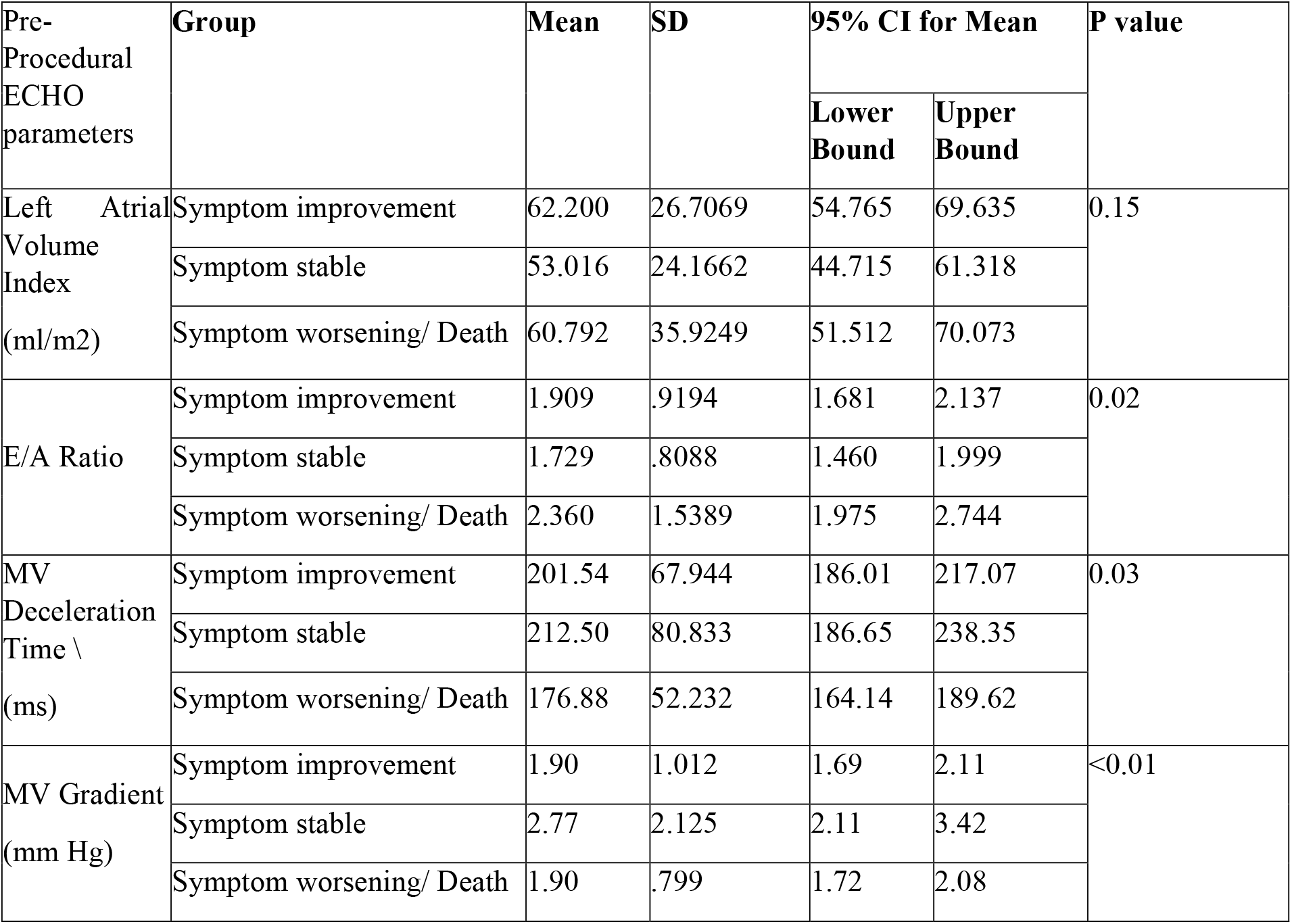
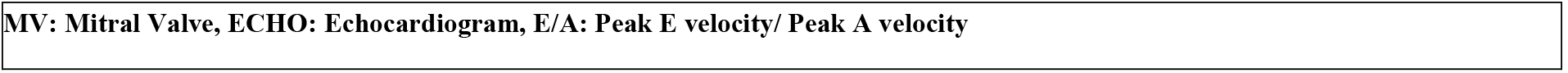
Pre-Procedural Echocardiographic parameters and variance across the groups.

| Pre-Procedural<br>ECHO<br>parameters | Group | Mean | SD | 95% CI for Mean |  | P value |
| --- | --- | --- | --- | --- | --- | --- |
|  |  |  |  | Lower<br>Bound | Upper<br>Bound |  |
| Left Atrial<br>Volume<br>Index<br>(ml/m2) | Symptom improvement | 62.200 | 26.7069 | 54.765 | 69.635 | 0.15 |
|  | Symptom stable | 53.016 | 24.1662 | 44.715 | 61.318 |  |
|  | Symptom worsening/ Death | 60.792 | 35.9249 | 51.512 | 70.073 |  |
| E/A Ratio | Symptom improvement | 1.909 | .9194 | 1.681 | 2.137 | 0.02 |
|  | Symptom stable | 1.729 | .8088 | 1.460 | 1.999 |  |
|  | Symptom worsening/ Death | 2.360 | 1.5389 | 1.975 | 2.744 |  |
| MV<br>Deceleration<br>Time \\<br>(ms) | Symptom improvement | 201.54 | 67.944 | 186.01 | 217.07 | 0.03 |
|  | Symptom stable | 212.50 | 80.833 | 186.65 | 238.35 |  |
|  | Symptom worsening/ Death | 176.88 | 52.232 | 164.14 | 189.62 |  |
| MV Gradient<br>(mm Hg) | Symptom improvement | 1.90 | 1.012 | 1.69 | 2.11 | <0.01 |
|  | Symptom stable | 2.77 | 2.125 | 2.11 | 3.42 |  |
|  | Symptom worsening/ Death | 1.90 | .799 | 1.72 | 2.08 |  |

MV: Mitral Valve, ECHO: Echocardiogram, E/A: Peak E velocity/ Peak A velocity

Results of the univariate regression analysis between pre procedural echocardiographic parameters and association with symptom worsening are summarized in Table 2.

**TABLE 2.** Association of Pre procedural Echocardiographic parameters with Symptom Worsening group.

| Pre-Procedural Echocardiographic Parameters | Odds Ratio of Association with Symptom Worsening Group | 95% Confidence Interval | P- value |
| --- | --- | --- | --- |
| E/A ratio | 1.51 | 1.10 - 2.06 | 0.010 |
| MV Deceleration time (ms) | 0.99 | 0.98 - 0.99 | 0.008 |
| MV gradient (mm Hg) | 0.822 | 0.63 - 1.07 | 0.148 |
| Left Atrial Volume Index (ml/m<sup>2</sup>) | 1.003 | 0.99-1.01 | 0.583 |
| MV: Mitral Valve, ECHO: Echocardiogram, E/A: Peak E velocity/ Peak A velocity |  |  |  |

Multivariate regression analysis was done comparing the E/A ratio in the pre procedural echocardiogram with the symptom worsening group. Results are summarized in Table 3.

**TABLE 3.** Multivariate Regression analysis for association of E/A ratio with Symptom Worsening.

| Pre-Procedural Echocardiographic Parameters | Odds Ratio of Association with Symptom Worsening Group | 95% Confidence Interval | P- value |
| --- | --- | --- | --- |
| E/A ratio | 1.61 | 1.13 - 2.29 | 0.008 |

E/A: Peak E velocity/ Peak A velocity

Area Under the Curve (AUC) was done between the pre procedural E/A ratio and symptom worsening group and depicted in Figure 1.

**FIGURE 1.**
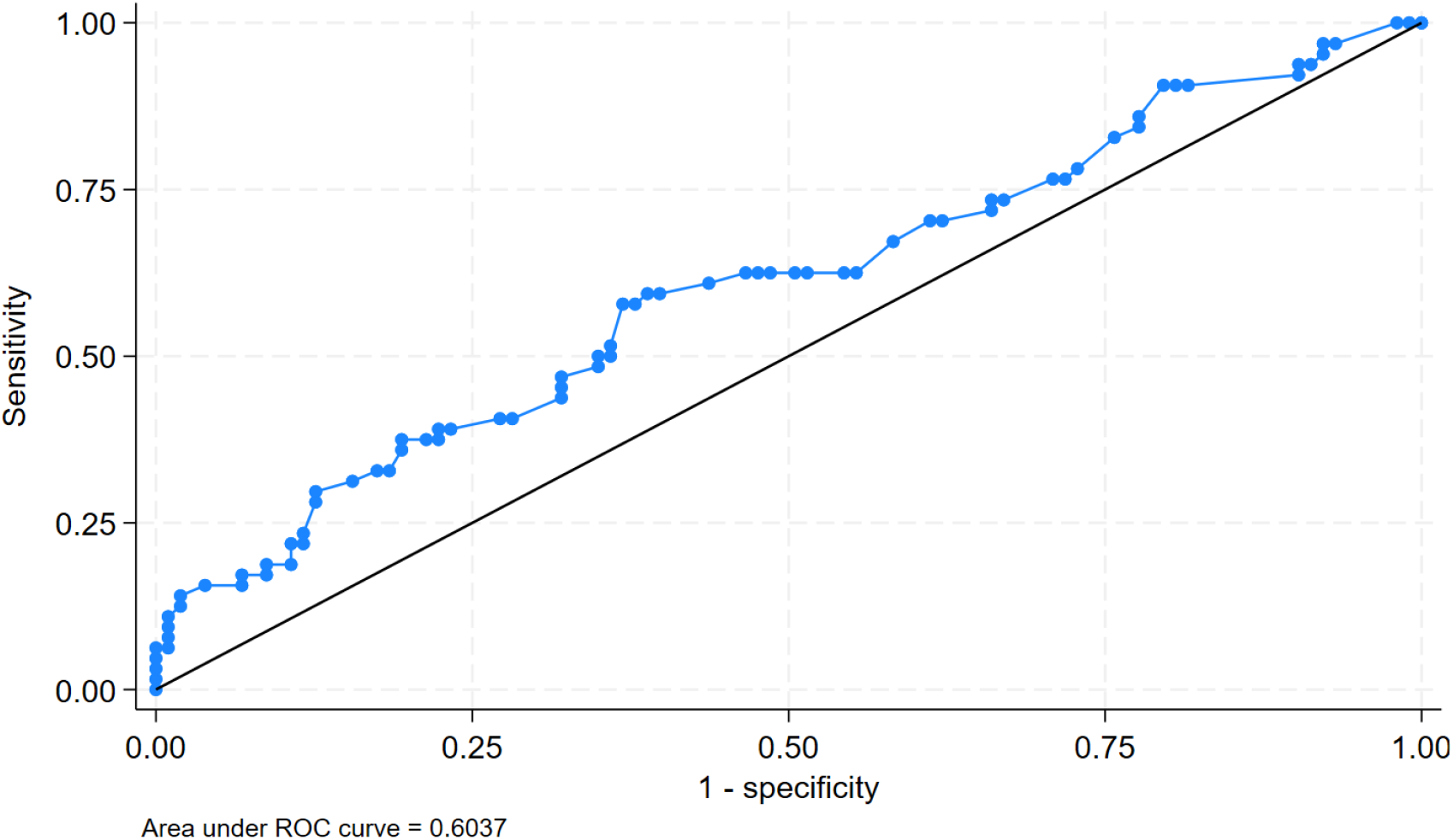
Area Under the Curve (AUC) analysis between Pre procedural E/A ratio and Symptom worsening.

**FIGURE 1a.**
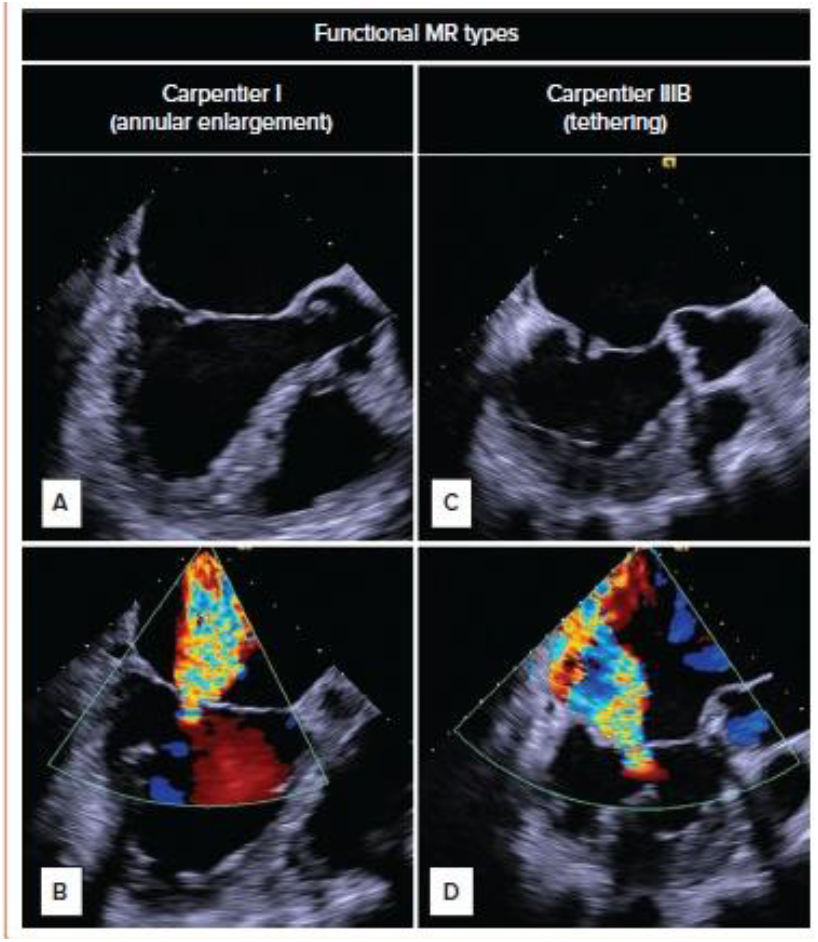
CARPENTIER CLASSIFICATION. **A:** Type1: Normal Leaflet Motion. **B**: Excessive leaflet motion, such as prolapse or flail leaflets, often due to papillary muscle rupture, chordae rupture, or redundant chordae. Seen more often in atrial fibrillation and non-ischemic cardiomyopathy **D:** Type IIIb: Restricted motion primarily during systole, commonly associated with left ventricular dilation, papillary muscle dysfunction, or tethering of the leaflets. Seen in Ischemic cardiomyopathy.

Based on Liu’s method, the optimal cutoff of E/A ratio to predict worsening status was 2.09, which provided a sensitivity of 58% and specificity of 63%. The area under the curve at this threshold was 0.60, indicating modest discriminative ability.

## DISCUSSION

The mitral valve apparatus is a dynamic structure with complex interactions surrounding its anatomy. Pathophysiologic mechanisms leading to FMR can be broadly divided into ischemic and non-ischemic causes [1]. Irrespective of the etiology, the primary driver is the lateral displacement of the annulus causing defective tethering forces. The decreased vertical tension created across the leaflets during systole can also be affected due to contractile dysfunction [11]. This more often leads to Carpentier IIIb functional regurgitation (Figure 1). Left atrial dilatation due to long standing atrial fibrillation could also lead to a pathology like FMR, presenting more often as a Carpentier type I functional regurgitation [11]. This explains how different pathophysiologic mechanisms can have an end common result of functional regurgitation across the mitral valve. Analyzing the studies and trials done on MitraClip^®^, meaningful patient outcomes occur when the regurgitation is the primary symptom driver as well as when the clip aids in reverse remodeling.

The role of diastolic function in the progression of secondary MR is not well studied. In subjects with ejection fraction less than 50%, E/A ratio is often used as a marker of Left ventricular diastolic filling (of LV relaxation and compliance). In MR, the elevated LA and LV pressures can lead to elevated pressure gradients, overestimating the MV peak E velocity and decreased LA function underestimating the MV A-velocity [12]. Hence E/A ratios are not used in the estimation of diastolic function in the presence of MR. While echocardiographic estimation of diastolic function can be difficult in the presence of MR, the role of impaired LA/LV relaxation and compliance can have significant effects on end diastolic pressures [13]. Irrespective of whether the etiology (ischemic versus non ischemic) of the FMR or the MR itself predominates, abnormal diastolic function can worsen the end diastolic pressures. The extent of diastolic dysfunction that could prohibit remodeling effects of mTEER has not been well documented in previous trials. AHA/ACC guidelines recommend a LVESD (Left Ventricular End Systolic Dimension) < 7 cm for mTEER in SMR, a threshold for remodeling beyond which the clip is unlikely to provide meaningful outcomes [2]. However, diastolic function parameters, that can play a significant role in determining end diastolic pressures, have not been traditionally used in patient selection criteria.

MV deceleration time and E/A ratio documented in the pre-procedural echocardiogram were the factors that had statistically significant variance across the symptom groups. MV deceleration time is inversely related to mitral regurgitation severity in severe PMR. This happens due to the rapid equilibration of pressures between the LA and the LV [14]. In a retrospective analysis of 234 patients that underwent undersized mitral annuloplasty ring, it was noted that an MV deceleration time of < 140 ms was predictive of worse outcomes [15]. In our analysis, the mean MV deceleration time in the symptom worsening group was 176.88 ms (164.14 - 189.62) and had a mean difference of -28.44 ms (95% CI: - 48.37 - -8.51). A lower MV deceleration time was associated with symptom worsening in the univariate analysis with an Odds Ratio of 0.99 (0.89 – 0.99), implying association between symptom worsening and a lower MV deceleration time. The association did not reach statistical significance in the multivariate regression analysis.

In our analysis, the pre-procedural E/A ratio had significant variance between the groups, with a p-value of 0.02 noted in the ANOVA. Running the chi-square test for pre-procedural E/A ratio, the symptom worsening group was found to have a significantly higher E/A ratio (mean difference 0.515, 95% CI: -0.88 - -0.14, p-value: 0.0067). In the multivariate regression analysis, a higher E/A ratio had association with the symptom worsening group, that reached statistical significance (aOR: 1.61 CI: 1.13 - 2.29). Age, Sex, baseline KCCQ scores, LVEDV, LVESV, EROA, PASP, history of atrial fibrillation, medication adherence and trans-mitral gradient were used in the regression analysis. Pre-procedural MV peak E velocity and MV deceleration time, despite having significant variance across outcome groups were avoided in the regression analysis due to collinearity with E/A ratio. In the ROC analysis, the higher E/A ratio had an AUC (area under the curve) of 0.603. The optimal cut off E/A ratio to predict symptom worsening was 2.094, which proved a sensitivity of 58% and specificity of 63%. A retrospective analysis involving 102 patients, a higher peak E velocity was found to have significant association with regurgitant fraction in FMR [16,17]. In this perspective, it can be argued that a higher E/A ratio could be reflective of the regurgitation severity. However, E/A ratio would still serve as a marker for LV relaxation and compliance, irrespective of the severity of regurgitation.

Left atrial volume index (LAVi) is yet another marker for the LV diastolic function and can be a critical marker of LA remodeling in MR. Elevated LAVi > 40 ml/m2 can be an independent predictive factor of worse outcomes in primary MR. Lavi also correlates with the severity of MR. As MR severity increases, so does LAVi, reflecting the chronic volume overload and subsequent LA dilation [18]. This relationship is evident in both chronic and acute MR settings, where LA distensibility and function are significantly impacted [19,20]. In clinical practice, LAVi is used to guide the management of primary MR. The American College of Cardiology/American Heart Association (ACC/AHA) guidelines recommend considering mitral valve surgery in asymptomatic patients with severe primary MR and LAVi > 60 mL/m^2^, even if left ventricular function is preserved, due to the high risk of adverse outcomes [21]. In our analysis (Table 1), no significant difference was observed in the pre procedural LAVi across the symptom groups. The trans-mitral gradient before mTEER implantation is another well studied parameter as a predictor for clip success. Studies have shown that appropriate mTEER placement with reduced EROA and trans-mitral gradient < 5 mmHg was a predictor of better outcomes [22]. Boerlage-van Dijk et al. found that the mean mitral valve gradient increased from 3.0 ± 1.6 mm Hg intraprocedural to 4.3 ± 2.2 mm Hg post procedurally, and further increased during exercise, indicating that operators should be cautious about the potential for elevated gradients post-procedure [23]. Additionally, Neuss et al. reported that a gradient exceeding 5 mm Hg post-implantation was associated with poorer long-term outcomes, including higher all-cause mortality and heart failure hospitalization [24]. In our analysis, the symptom groups did not differ statistically in terms of the pre or post procedural mean mitral valve gradient or the increase in gradient after mTEER implantation (Table 2).

## CONCLUSIONS

E/A ratio, MV deceleration time and trans-mitral gradient obtained in the pre-procedural echocardiogram had significant variance across the outcome groups. Patients in the symptom worsening group had a lower mean MV deceleration time, and a lower MV deceleration time was associated with symptom worsening in the univariate regression analysis. However, the association between MV deceleration time and symptom worsening/death lost significance in the multivariate regression analysis. A higher E/A ratio was associated with symptom worsening/death in the multivariate regression analysis (Adjusted OR: 1.61 CI: 1.13 - 2.29, p value: 0.008). ROC analysis between E/A ratio and symptom worsening/death had an AUC of 0.603, corresponding to a modest degree of predictability compared to random guessing.

## Data Availability

RAW data added as supplementary file

## LIMITATIONS OF THE STUDY

The study was designed in a retrospective longitudinal cross-sectional format. The results must be interpreted as a hypothesis generation. Further randomized studies need to be done to assess the causal association of diastology and outcomes after mTEER.

